# COVID-19 Serology in New York State Using a Multiplex Microsphere Immunoassay

**DOI:** 10.1101/2021.05.12.21257125

**Authors:** Danielle T. Hunt, Jennifer L. Yates, Karen E. Kulas, Kyle Carson, Theresa Lamson, Valerie Demarest, Andrea Furuya, Kelly Howard, Mary Marchewka, Randy Stone, Heidi Tucker, Casey Warszycki, Jim Yee, He S. Yang, Sabrina Racine-Brzostek, Zhen Zhao, Monir Ejemel, Qi Li, Yang Wang, Sebastian Fernando, Francesca La Carpia, Eldad A. Hod, Kathleen A. McDonough, William T. Lee

**Affiliations:** Division of Infectious Diseases, Wadsworth Center, New York State Department of Health, Albany, NY, 12208 USA; Department of Biomedical Sciences, The School of Public Health, The University at Albany, Albany, NY, 12222 USA; Department of Pathology and Laboratory Medicine, Weill Cornell Medicine, New York, NY, USA; NewYork-Presbyterian Hospital/Weill Cornell Medical Campus, New York, NY, USA; MassBiologics of the University of Massachusetts Medical School, Boston, MA, 9 02126 USA; Department of Pathology and Cell Biology, Columbia University Irving Medical Center, New York, NY, USA

**Author notes:** Address Correspondence to: Dr. William T. Lee, David Axelrod Institute, The Wadsworth Center/NYSDOH, 120 New Scotland Avenue.

## Abstract

The emergence of SARS-CoV-2, leading to COVID-19, necessitated the development of new molecular and serological tests. Here, we describe a multiplexed serological assay developed as the global pandemic moved into New York State in the spring of 2020. The original microsphere immunoassay used a target antigen from the SARS-CoV-1 virus responsible for the 2003 SARS outbreak, but evolved to incorporate multiple SARS-CoV-2 protein antigens (nucleocapsid, spike and spike domains, spike and nucleocapsid proteins from seasonal human coronaviruses). Besides being highly versatile due to multiplex capabilities, the assay was highly specific and sensitive and adaptable to measuring both total antibodies and antibody isotypes. While determining the assay performance characteristics, we were able to identify antibody production patterns (e.g., kinetics of isotypes, individual variations) for total antibodies and individual antibody classes. Overall, the results provide insights into the laboratory response to new serology needs, and how the evolution and fine-tuning of a serology assay helped contribute to a better understanding of the antibody response to SARS-CoV-2.

## 1. Introduction

The coronavirus disease 2019 (COVID-19) (1), caused by the Severe Acute Respiratory Syndrome Coronavirus 2 (SARS-CoV-2)(2), is historically one of the most widespread and devastating pandemics with severe health, economic, and social consequences (3, 4). As a newly emergent virus, knowledge of the resulting disease courses, host response characteristics, and the development of diagnostic tests arose only as the pandemic progressed. Still, a strong framework of information was already present from prior studies on other disease-causing coronaviruses, including circulating human endemic coronaviruses (HuCOV), Middle East Respiratory Syndrome virus, and the closely related SARS-CoV-1 (reviewed in (5, 6)).

Within the past year there has been an enormous growth in the number of laboratory assays designed to diagnose SARS-CoV-2 infection. These primarily rely on molecular analyses for the detection of viral RNA, and, more recently, serological tests to detect viral antigens and host antibodies (Abs) for the determination of prior exposure to SARS-CoV-2. In particular, there are a wide variety of serology tests with different performance characteristics, different target antigens, and different readouts (7). These tests have clinical, surveillance, and research applications, including information about the COVID-19 disease process and host immune response.

The impetus for this study was an exigent need for early development of molecular and serological assays to meet the COVID-19 outbreak in New York State (NYS). As the laboratory arm of the New York State Department of Health, the Wadsworth Center was heavily engaged in assay development and early testing. The Diagnostic Immunology Laboratory at the Wadsworth Center (DIL) initiated an effort to develop a serological assay to identify persons exposed to the novel coronavirus. We developed an initial microsphere immunoassay (MIA) using a Luminex® platform. We determined performance characteristics and implemented the assay for COVID-19 serology testing of sera from NYS residents. This platform is very adaptable which allows the inclusion or replacement of several different target molecules. As described in this report, over the course of the outbreak, we frequently assessed the testing data and made changes as needed, which also helped to improve our understanding of the SARS-CoV-2 humoral response to infection.

## 2. Results

### 2.1 Establishment of the MIA

The NYS SARS-CoV-2 MIA was initially conceived during the early phase of the Wuhan COVID-19 outbreak (1). As no SARS CoV-2 proteins were available to us at the beginning of February 2020, the first generation of the assay used the SARS-CoV-1 nucleocapsid (N) protein as the target antigen (8). The SARS-CoV-1 N protein, prepared at the Wadsworth Center in 2003 for use in the original SARS outbreak (9), was coupled to microspheres and tested using a guinea pig anti-serum reactive with SARS-CoV-1. The antiserum showed high binding (Figure 1A), while serum from a nonimmune guinea pig did not react. In later experiments employing SARS-CoV-2 antigens and COVID-19 sera, the same guinea pig anti-SARS-CoV-1 serum showed nearly equivalent binding to the SARS CoV-2 N protein (Figure 1A). Likewise, COVID-19 patient sera were comparably reactive with both SARS-CoV-1 and SARS-CoV-2 N proteins (Figure 1B) and, MIAs using either protein as the target antigen behaved equivalently (Figure 1C). The high degree of cross-reactivity between the two N antigens contrasted with our later observations of differential antibody binding to the SARS-CoV-1 and SARS-CoV-2 Spike proteins, as we observed little binding of the SARS-CoV-1 guinea pig serum to the SARS-CoV-2 spike proteins (Figure 1D) and, conversely, reduced binding of COVID-19 sera to the SARS-CoV-1 spike protein (Figure 1E and F).

**Figure 1.**
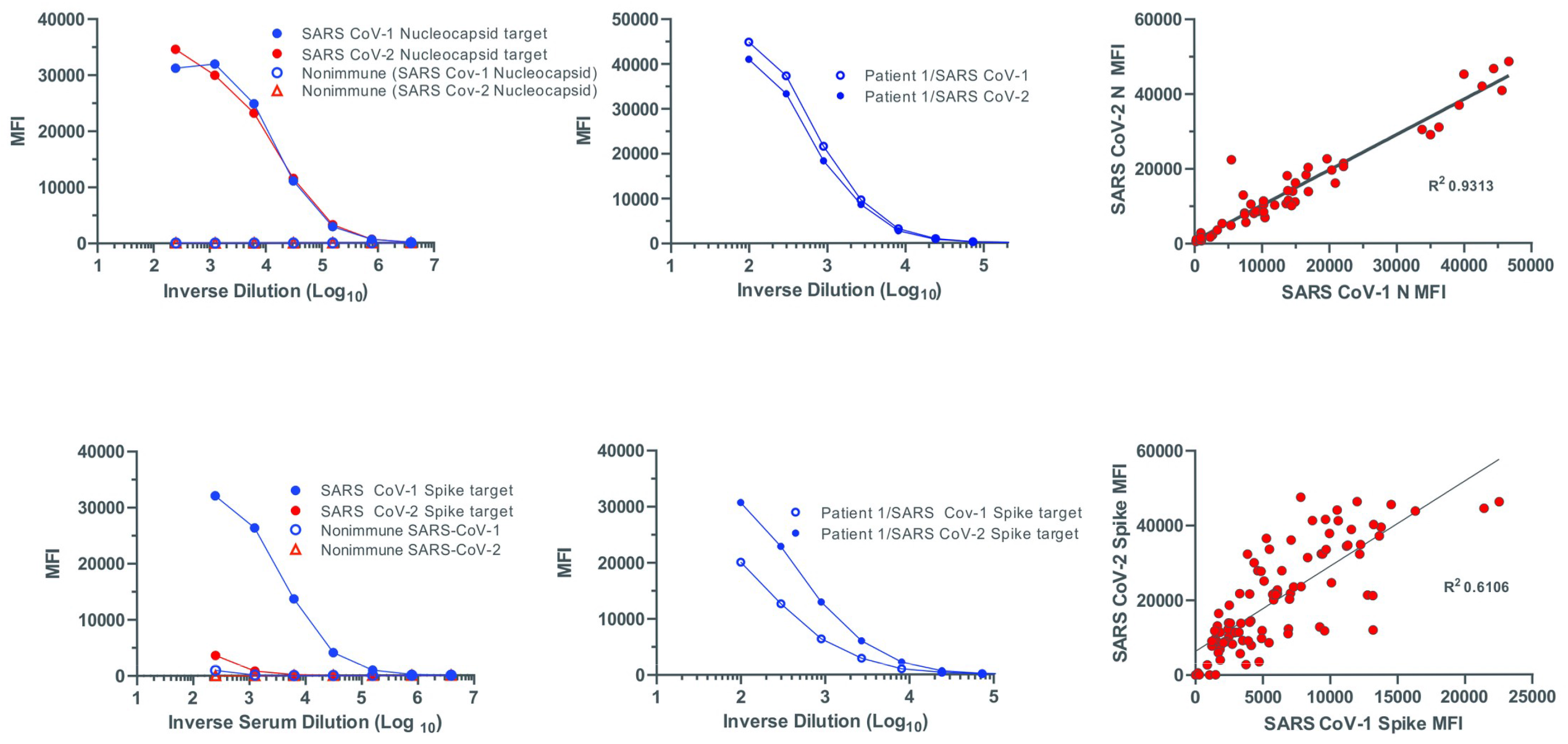
Cross-reactivity between SARS-1 and SARS CoV-2 Nucleocapsid and Spike antigens. Sera from nonimmune or SARS-CoV-1-immunized guinea pigs (*A,D*), a COVID-19 convalescent individual (*B,E*) or a panel of sera from COVID convalescent individuals (*C,F*) were diluted and measured in a MIA for total binding antibodies against the Nucleocapsid (*top row*) and Spike antigens (*bottom row*) from either SARS-1 or SARS-CoV-2. Binding activity is indicated by the median fluorescence intensities (MFI).

### 2.2 Specificity of the MIA

Assay cutoffs are established as an MFI threshold to determine specimen reactivity. A panel of 92 serum specimens from healthy donors, collected in 2009, were tested for total antibody produced against SARS-CoV-1 N-protein and the cutoff was defined as the mean median fluorescence intensities (MFI) plus three standard deviations (SD) (Figure 2A). An additional 164 specimens were tested, and, of the total of 256, only 7% of the specimens (93% specificity) were reactive (data not shown). Considering that for endemic seasonal coronaviruses (HCoV-229E, -NL63, -OC43, and -HKU1) there is an estimated rate of up to 15% infections per year in a geographically close region (10), we reasoned that there was little impact in the assay by antibodies to endemic coronaviruses. We next tested a panel of 30 specimens collected 10-20 days post onset of symptoms from patients with diagnosed <u>non-COVID-19</u> respiratory illnesses, many of whom had documented positive PCR tests for specific pathogens. Several of the specimens in this category were reactive with the SARS-CoV-1 N protein in the first-generation MIA, lowering the assay specificity to 67% (Figure 2B). The most notable cross-reactive specimens were those from patients with RT-PCR-confirmed recent exposure to the endemic coronavirus HCoV-NL-63 coronavirus, for which half of the tested specimens had reactivity. The overall MFIs of the cross-reactive specimens, including the reactive HCoV-NL-63 specimens, were lower than the majority of signals from COVID-19 specimens (Figure 2C); hence, to allow for a more specific test, even at the expense of sensitivity, we raised the cutoff for the assay to 6 SD above the mean MFI of the healthy donor sera. At this level, when measuring total antibodies, the specificity was 99.2% (N only). A second-generation MIA was later implemented and used the SARS-Cov-2 N antigen and, to better capture the breadth of responses, the SARS-CoV-2 receptor-binding domain (RBD) of the spike protein. The overall specificity of this assay was 98.4% (N plus RBD) (Table 1). Including sera containing Abs to known pathogens or autoantigens did not reduce the specificity (Table 1). For clinical test reporting, the specimens with MFI values that fell between 3 SD and 6 SD of the mean MFI of the normal serum specimens were listed as “Indeterminate” (Figure 2C). In the second-generation MIA, that included the SARS-CoV-2 RBD, when the two target antigens gave different results, the overall result was based on the most positive of either antigen (e.g., indeterminate plus reactive was reported as reactive). Subsequent analyses of the pre-COVID blood donors showed that there were abundant antibodies to both the N and spike antigens of the seasonal coronaviruses (Supplemental Figure 1), strongly suggesting that <u>non-acute</u> antibodies to the seasonal coronaviruses did not cross-react with the SARS-CoV-2 antigens in the MIA.

**Table 1.** Determination of MIA Specificity. Sera collections were used to evaluate total antibody (Ig) or isotype-specific binding to either the SARS-CoV-2 N protein or the RBD of the spike protein in a multiplexed MIA: (top) 182-257 serum samples collected in 2009 from healthy blood donors (American Red Cross in Syracuse, NY, and New York Blood Center in New York City, NY), representing pre-COVID samples; (bottom) 78 serum samples with known antibody reactivity to a diverse group of viral pathogens (Chikungunya, dengue, HCV, HIV, measles, mumps, rubella, VZV, West Nile virus, herpes simplex, Zika virus, enteroviruses, HBV, cytomegalovirus, Epstein Barr, Eastern Equine Encephalitis, and Yellow Fever viruses), as well as sera with Antinuclear antibodies and Rheumatoid Factor. Shown are the numbers (percentages) of MIA negative/total specimens and the 95% Confidence Intervals. “Negative” means non-reactive to both target antigens in the specimen; “indeterminate” means indeterminant to one target antigen and indeterminant or non-reactive to the other target antigen in the specimen. For this analysis, indeterminates and negatives are counted together, using a threshold of 6 SD above the mean MFI of 92 normal sera to determine positivity.

|  | **Non-Reactivity (95%CI) in the MIA (N plus RBD)** |  |  |  |
| --- | --- | --- | --- | --- |
|  | *Ig* | *IgM* | *IgG* | *IgA* |
| **Blood Donors** | 240/244 (98.4%)<br>(95.9-99.4) | 248/257 (96.5%)<br>(93.5-98.2) | 254/257 (98.8%)<br>(96.6-99.7) | 182/186 (97.8%)<br>(94.6-99.2) |
| **Diverse Group of Analytes** | 77/78 (98.7%)<br>(93.1-99.9) | 74/78 (94.9%)<br>(87.5-98.0) | 76/78 (97.4%)<br>(91.1-99.5) | 77/78 (98.7%)<br>(93.1-99.9) |

**Figure 2.**
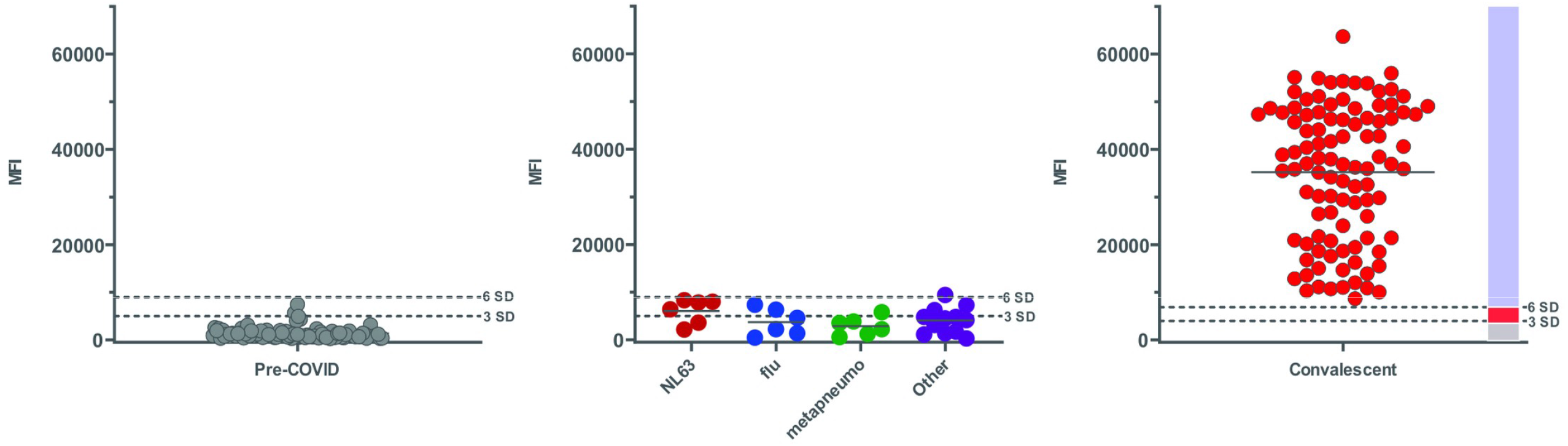
Determination of NYS SARS MIA specificity. (*A*) Serum specimens collected in 2009 from healthy blood donors (American Red Cross in Syracuse, NY, and New York Blood Center in New York City, NY), representing pre-COVID samples or (*B*) specimens collected in 2020 from hospitalized SARS-CoV-2 RT-PCR negative individuals with molecular determination of infection with non-SARS-CoV-2 respiratory viruses; or (*C*) specimens collected in 2020 from COVID-19 convalescent individuals were assessed using the NYS SARS MIA (SARS-1 N antigen target) for total antibody reactivity. Shown are results of individual samples. On the far right is a bar showing the reported interpretative results of (*blue*) Reactive, (*Red*) Indeterminate or (*gray*) Nonreactive.

### 2.3 Sensitivity of the MIA

A broad examination of COVID-19 specimens showed that a greater number were reactive as time progressed (Figure 3). Examination of total antibody reactive in the MIA, regardless of using only the N antigen (Figure 3) or both the N and RBD proteins as target antigens (Table 2), showed that the sensitivity of the assay slowly increased with increased time between symptom onset and testing, with a sensitivity of 96.4% 26-30 days following symptom onset (Table 2). Notably, at the time the initial MIA (based upon SARS-CoV-1 N antigen) was submitted to the Food and Drug Administration (FDA) for Emergency Use Authorization (EUA), the maximum days of post symptom onset available to us was 25 days, in which only 88% of the specimens had N-specific antibodies detected by the MIA above the 6SD cutoff (EUA: https://www.fda.gov/media/137541/download). Hence, the initial (FDA) published sensitivity of our N antigen MIA was 88%. Subsequent testing of specimens with >30 days post symptom onset, showed that the assay could detect a much higher percentage of positive specimens. For example, an analysis of 8659 COVID-19 convalescent sera (collected 30-50 days post symptom onset) showed that the assay had a 92.2% (N antigen-reactive) sensitivity using the 6 SD cutoff, compared to 95.9% using 3 SD (Table 3). This study also showed the impact of adding additional antigens, such as the RBD, into the MIA. Comparison of separate reactivity to the N and the RBD proteins within the same specimens showed that, while most sera contained antibodies reactive with both antigens, several had reactivity only to the N protein (4.2%, Table 3). Reactivity to RBD protein without reactivity to N did occur but at a lower frequency (<1.2% of reactive specimens) (Table 3). When examining individual specimens, although more often a high N reactivity coincided with high RBD reactivity, many specimens had high Ab levels to one antigen but lower levels to the other (data not shown). Our recent studies showed that by adding the complete spike antigen, or by replacing the RBD with the spike antigen, the sensitivity of the MIA could be even further increased (Supplementary Table 1). However, either with just the RBD or with the spike antigen, the MIA performs comparably to other COVID-19 serology assays. In a study looking at 500 sera, including 100 pre-COVID sera, the MIA had similar or better sensitivity and specificity as three highly used commercial laboratory tests performed on the Ortho Vitros, Biomerieux Vidas, or Abbot Architect platforms (Supplementary Table 1).

**Table 2.** Determination of MIA Sensitivity. A total of 93 serum specimens from individuals with SARS-CoV-2 infections, confirmed using RT-PCR, and reported symptom onset dates were assessed for total antibodies to the SARS-CoV-2 Nucleocapsid antigen and the RBD. A positive specimen is ≥6 SD above the cutoff for either one of the target antigens; an indeterminate is ≥3 SD above the cutoff for either or both of the target antigens; a negative is a sample with no reactivity to either target antigen. Sensitivity at different period post symptom onset is shown on the right as the number of MIA-positive specimens/total specimens tested.

| Total Antibody binding to N plus RBD |  |  |  |  |  |  |
| --- | --- | --- | --- | --- | --- | --- |
| Days from onset | Samples tested (N) | RT-PCR result | NY SARS-CoV MIA Result as Compared to PCR |  |  | Sensitivity (95%CI) |
|  |  |  | Positive | Indeterminate | Negative |  |
| 7 – 15 | 48 | Pos | 23 | 7 | 18 | **23/48**<br>**47.9% (34.47% - 61.67%)** |
| 16 – 25 | 17 | Pos | 14 | 0 | 3 | **14/17**<br>**82.4% (58.97% - 93.81%)** |
| 26 – 30 | 28 | Pos | 27 | 1 | 0 | **27/28**<br>**96.4% (82.29% - 99.82%)** |
| **Total** | **93** |  |  |  |  |  |

**Table 3.** Reactivity to the SARS-COV-2 N and RBD antigens. 8659 serum specimens from convalescent COVID-19 donors who were at least 14 days removed from symptom onset were tested using the MIA for total antibody reactivity to the SARS-COV-2 and RBD antigens. A reactive specimen is ≥6 SD above the cutoff; an indeterminate is ≥3 SD above the cutoff. The bottom four rows indicate assay reported results calculated to show positivity at 3SD or 6 SD thresholds and shows the effect of using the single N target Ag versus both target Ags. * Threshold used for reporting “Reactive” specimens in diagnostic assays. **One or both target Ags reactive.

| Convalescent Serum Specimens | Number | Percentage |
| --- | --- | --- |
| **Total Tested** | **8659** | **100** |
| N and RBD<br>(Both Reactive or Indeterminate in the same sample) | 7980 | 92.2 |
| N (Reactive or Indeterminate) with RBD (Nonreactive) | 365 | 4.2 |
| RBD (Reactive or Indeterminate) with N (Nonreactive) | 101 | 1.2 |
| Both antigens Nonreactive | 238 | 2.7 |
| One or both antigen Indeterminate (*clinical\**) | 192 | 2.2 |
| Using N only, Positive @6 SD (*clinical\**) | 7986 | 92.2 |
| Using N only, Positive @3 SD | 8304 | 95.9 |
| Using N and RBD**, Positive @6 SD (*clinical\**) | 8219 | 94.9 |
| Using N and RBD**, Positive @3 SD | 8411 | 97.1 |

**Figure 3.**
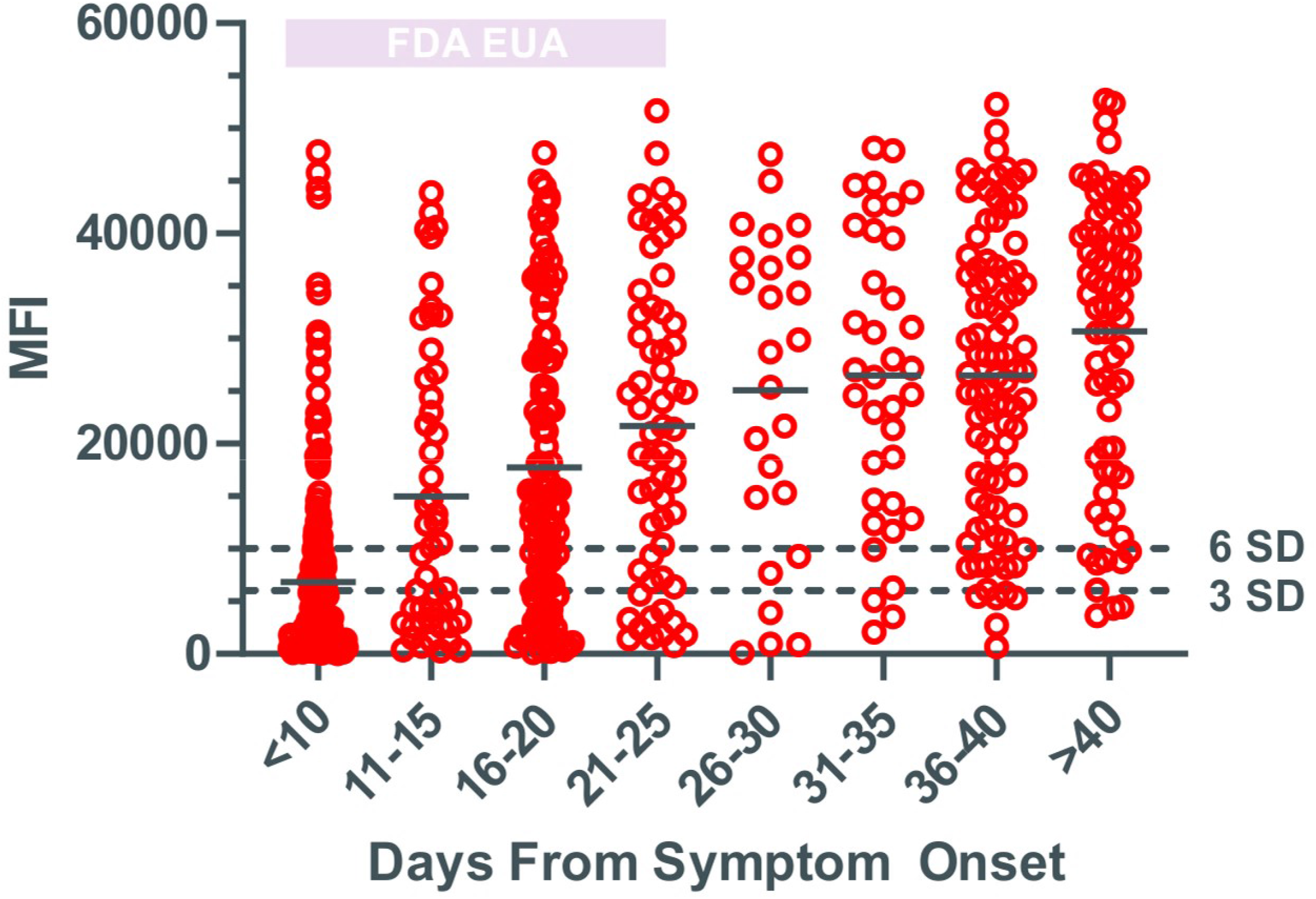
Detection of N antigen reactivity by MIA. Serum specimens from individuals with SARS-CoV-2 infections, confirmed using RT-PCR, and reported symptom onset dates were assessed for total antibodies to the SARS-1 N antigen. Each point represents the reactivity of a single individual with the mean MFI of the group at each time point indicated by a black bar. The gray bar at the top left indicates the data accumulated for the initial submission of the MIA for FDA EUA with assay sensitivity determined as the percentage of specimens reactive at ≥6 SD above the cutoff.

Although the primary MIA developed and used for clinical testing by the DIL measured total antibody, the assay proved readily adaptable to both greater multiplexing of antigens and to the measurement of individual Ab isotypes. For example, the study in Supplementary Figure 1 shows the multiplexing capability with simultaneous measurement of Abs to the N and spike antigens of the four seasonal coronaviruses and the same antigens of SARS-CoV-2 (10-plex). The study shown in Supplementary Table 1 and 2 shows the measurement of individual Ab isotypes. The isotype MIAs exhibited the same specificity as the parent total Ab MIA (Table 1) and exhibited good sensitivity which increased with time after post-symptom onset (Supplementary Table 2).

### 2.4 Isotype Switching in Response to SARS-CoV-2

Figure 4A demonstrates that there is a wide range in individual production of total Ab made to both the N and RBD antigens and shows that, even early on, some individuals were highly reactive in the MIA. We more deeply investigated the onset of Ab production by determining the kinetics and distribution of antibody classes. We assessed sera from individuals at different times from onset of COVID-19 symptoms using the isotype-specific version of the MIA while examining the responses to multiple SARS-CoV-2 antigens. For these studies, we not only looked at reactivity to the RBD of the spike antigen, but also the entire spike and its two major domains: the amino-terminal S1 subunit of the spike, which contains the RBD, and the carboxy-terminal S2 subunit. Although most of the anti-spike Abs appear to be directed toward S1 in most individuals, the S2 is also immunogenic (Figure 4B and Supplementary Figure 2). The higher reactivity towards the S1 domain, as compared to the RBD, suggests that of the S1-reactive Abs, the RBD appears to be one, but not the only, focus of reactivity as measured by this assay. Examination of antibody classes directed against the panel of SARS-CoV-2 target Ags showed that smaller amounts of IgM antibodies were directed toward the N antigen and S2 domain of the spike protein, as compared to the RBD and S1 antigens (Figure 5 and Supplementary Figure 3). This was reflected both in the proportion of individuals making Abs and the abundance of Abs. This contrasts with IgG production where we observed similar production against all the tested antigens. Likewise, we observed IgA production with all the tested antigens, with, perhaps, slightly better IgA responses to the S1/RBD region of the spike (Supplementary Figure 3).

**Figure 4.**
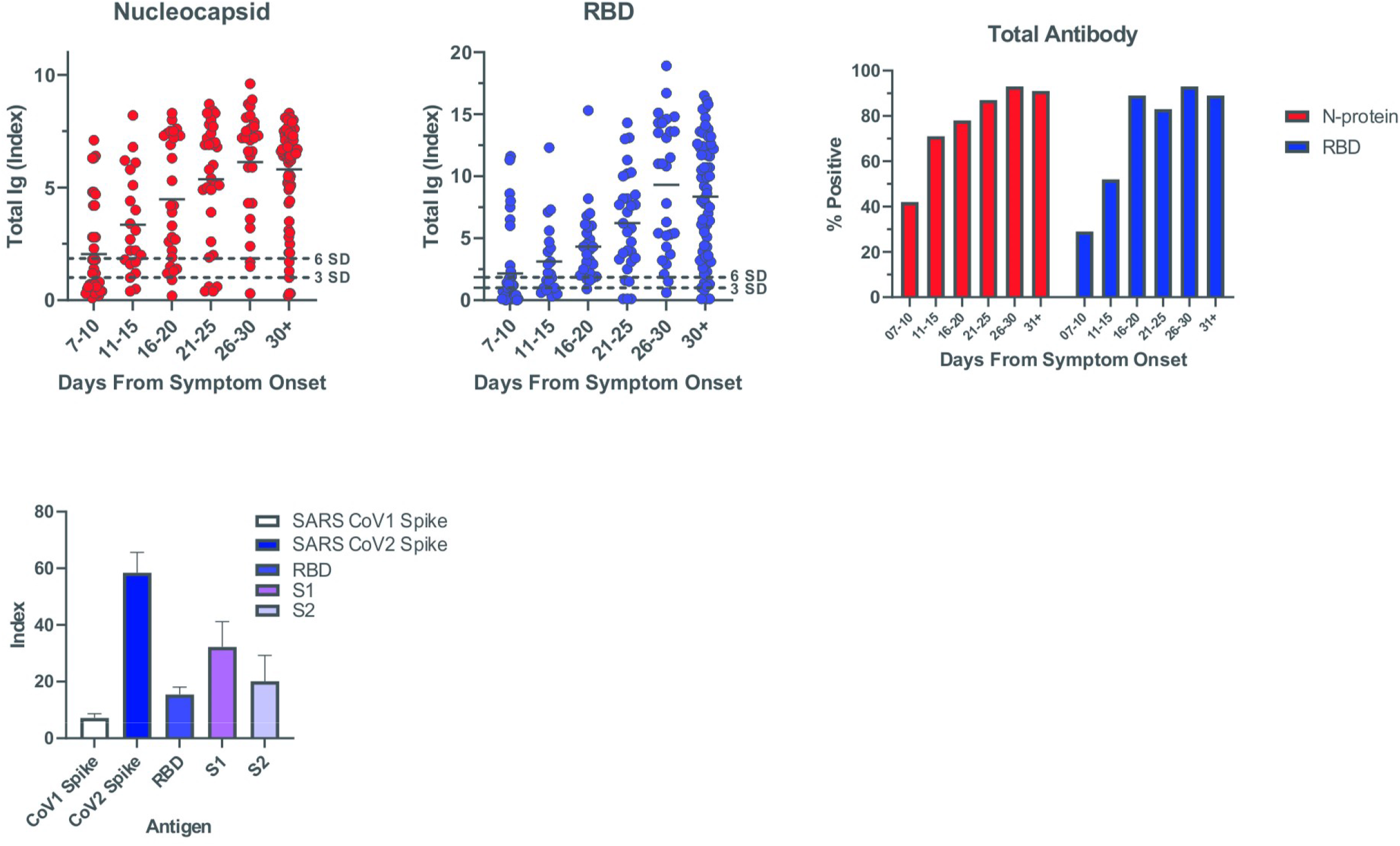
Reactivity to different SARS-CoV-2 target antigens as determined by MIA. Serum specimens from individuals with SARS-CoV-2 infections, confirmed using RT-PCR, and reported symptom onset dates were assessed for total antibodies to the SARS-CoV-1 spike antigen or different SARS-CoV-2 target antigens (Nucleocapsid, complete spike, and separate spike components (RBD, S1 subunit, S2 subunit). Reactivity is expressed as an Index value where a value of 1.0 is 3 SD above the cutoff for the individual target antigen and positivity being the Index value that is ≥ 6 SD above the cutoff for the individual target antigen. (*A*) The reactivity in the same sera to Nucleocapsid and RBD antigens at different times after onset of symptoms shown for individuals or as a percent of the total sera (far right panel). (*B*) Relative binding to multiplexed antigens in the sera of 10 COVID-19 convalescent individuals (means + SD).

**Figure 5.**
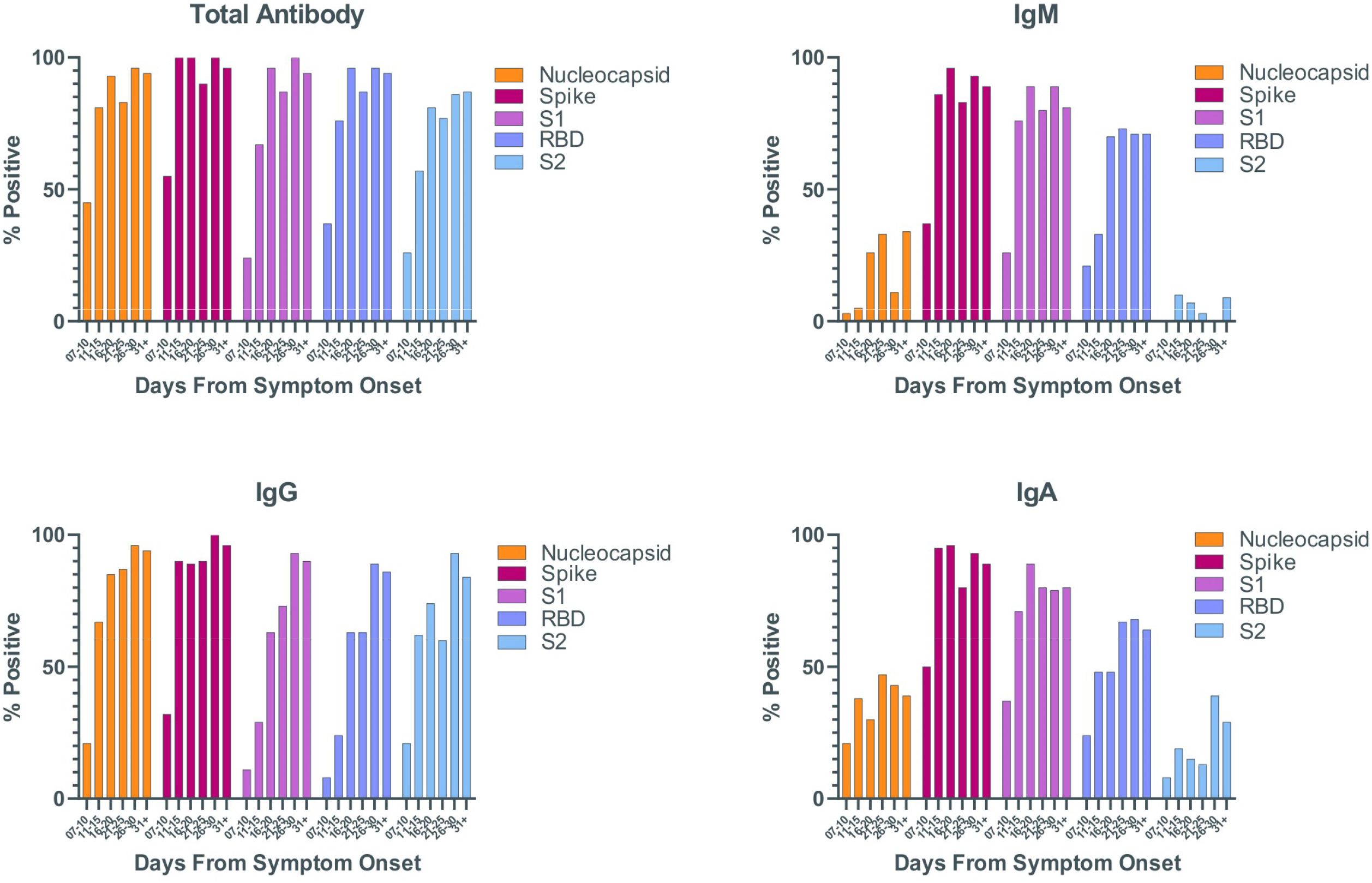
Production of different antibody classes reactive with SARS-CoV-2 target antigens. Serum specimens from individuals with SARS-CoV-2 infections, confirmed using RT-PCR, and reported symptom onset dates were assessed for total antibody, IgM, IgG, and IgA antibodies to different SARS-CoV-2 target antigens (Nucleocapsid, full spike, and separate spike components (RBD, S1 subunit, S2 subunit). Shown are the percentages of positive specimens for each class to the individual target antigens.

There is clearly individual variation, with some specimens having copious amounts of antibodies and early production of antibodies. However, overall there was a progressive increase of total antibodies that did not peak until after 40 days post symptom onset (Figure 3). In contrast with canonical kinetics of secreted Ab isotypes, we found that IgM did not significantly precede the production of IgG; indeed, a higher percentage of IgG positive specimens were found at the earlier times after symptom onset, as compared to IgM (Figure 5). The rapid rise of IgG can be more clearly seen when looking at serial specimens from the same individual (Figure 6). We also examined the production of IgG subclasses. We found a robust IgG1 response to all the target antigens that was induced relatively early in the infection (Figure 7, Supplementary Figure 4). IgG3 production was also robust, albeit slightly delayed. In contrast, only a small percentage of specimens contained detectable IgG2 and IgG4. Detectable IgG4 production appeared to be restricted to the nucleocapsid antigen (Figure 7, Supplementary Figure 4).

**Figure 6.**
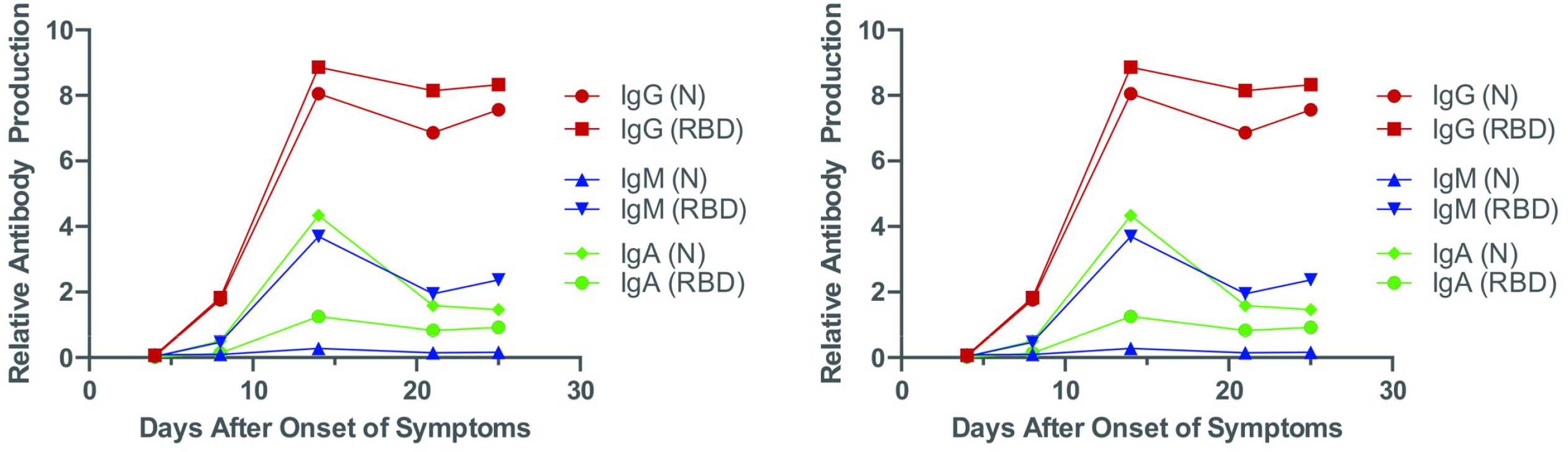
Kinetics of different antibody class production in COVID-19 individuals. Serum specimens from two individuals hospitalized with SARS-CoV-2 infections were collected on different days after reported symptom onset dates and were assessed for IgG1, IgG2, IgG3, and antibodies to different SARS-CoV-2 target antigens (Nucleocapsid, and separate spike components (RBD, S1 subunit, S2 subunit). Shown are the percentages of positive specimens for each class to the individual target antigens.

**Figure 7.**
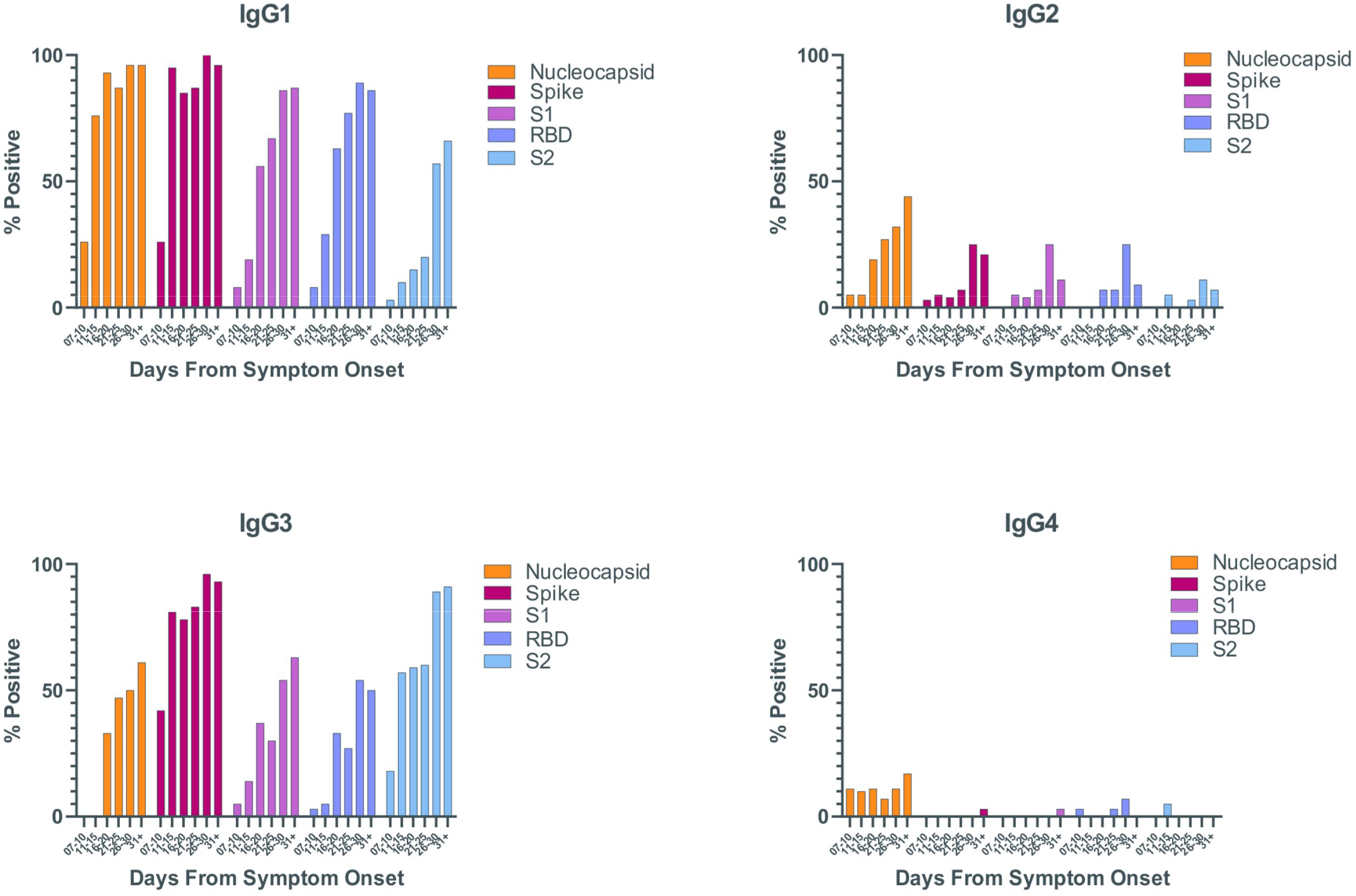
Production of different IgG subclasses reactive with SARS-CoV-2 target antigens in the MIA. Serum specimens from individuals with SARS-CoV-2 infections, confirmed using RT-PCR, and reported symptom onset dates were assessed for IgG1, IgG2, IgG3, and IgG4 antibodies to SARS-CoV-2 nucleocapsid, spike antigen and spike components: RBD, S1, and S2 target antigens. Shown are the percentages of positive specimens for each class to the individual target antigens

## 3. Discussion

This report describes the overall experience of the Wadsworth Center’s approach in developing serology testing to determine prior SARS-CoV-2 virus exposure. The test was developed to address an emergency public health need in New York, which initially had the highest number of COVID-19 cases in the US (11). While not the first US clinical COVID-19 serology assay to receive FDA EUA, ours was the first microsphere-based assay to receive EUA ((12) and EUA: https://www.fda.gov/media/137541/download). Other MIAs have also been described for use in research studies (e.g., (13, 14)) and, for clinical testing, Luminex Corp subsequently received EUA for its MIA (12). It was reassuring that, despite our initial MIA being based on the SARS-CoV-1 N antigen, it showed good result agreement with the first EUA ELISA, based on the SARS-CoV-2 RBD target antigen, developed by the Krammer laboratory (15, 16). However, this should not have been surprising, given the high degree of antigenic cross-reactivity between the N antigens of the two viruses. That cross-reactivity also reflects the degree of amino acid homology (90%) (8), as does the lower cross-reactivity observed between the spike antigens of SARS-CoV-1 and SARS-CoV-2 (76% amino acid homology) (8).

Amino acid homology between the N or spike antigens of SARS-CoV-2 and their counterparts of the four seasonal coronaviruses is <50% (17), and, likewise, none of the common coronaviruses interfere with the SARS-CoV-2 MIA. It is interesting however, that, although normal sera contain abundant antibodies to the common coronaviruses, we only found significant reactivity when assessing sera from individuals recently infected with respiratory viruses, including common coronaviruses. Whether this reflects induced cross-reactive antigens or is a consequence of inflammation associated with the acute infections, is currently unclear. It is notable that increases in antibodies reactive with common coronaviruses also occur in acute COVID-19 infections. Acute respiratory infection sera are not usually included when determining assay specificity, but, if generalizable to other assays, the interference with the COVID response should be considered when using highly sensitive tests when both HuCoV’s and influenza infections are prevalent. With respect to our SARS-CoV-2 MIA, these sera prompted us to raise the assay threshold and sacrifice some sensitivity (ultimately, 92.2% versus 95.9% using only the N antigens; 94.9% versus 97.1% using multiplexed N and RBD antigens). The sensitivity is improved over that of the original assay, due to not only the addition of the N antigen, but also the testing of specimen that had been collected at a later time point after symptom onset. As measured by our MIA, the COVID antibody response over a broad group of individuals was slower than expected, with the highest percentages of Ab-positive individuals occurring after 21 days. Further, a small number of individuals did not appear to make substantial amounts of antibodies in response to infection. Even at the 3 SD level, 2.7% of the specimens were negative in this study. It is likely that these are individuals who truly failed to mount a large Ab response to the virus. In an examination of >27,000 convalescent sera, we identified donors who had been tested up to 8 times over a 6-month period, with certain individuals having consistently undetectable Ab levels in the MIA (unpublished observations). Given that these represent convalescent individuals, it appears that in some instances, COVID-19 can be resolved with minimal antibody production, presumably solely through cell-mediated immunity. This conclusion would be consistent with studies showing that, although neutralizing antibody plays a predominant role, T cells can contribute to protection when antibody levels are suboptimal (18-20).

Since the pandemic began, numerous serology tests have been developed and implemented for both research and clinical testing. To date, over 60 individual serology assays have received EUA (12). These tests measure antibodies to a variety of target antigens and generally measure total antibody or IgG (with or without separate detection of IgM). Most tests identify antibodies reactive with <u>either</u> the N antigen or a version of the spike protein (RBD, S1, or full trimeric spike). However, the abundance of antibodies to the N protein or spike protein are independent of each other and an individual serum might have strong reactivity to one antigen with poor or absent reactivity to the other. This may make result comparisons between different assays difficult. The detection of only one or the other Ag reactivity is an important consideration when comparing performance of assays that use different target antigens. An advantage of the MIA is that antigens can be easily and routinely multiplexed. For example, we show here simultaneous measurements of antibodies reactive with the N, S1, S2, RBD, trimeric spike, and seasonal coronavirus N plus spike antigens in a single specimen. Examination of specific antibody responses to each antigen may also be important for identifying severity parameters of COVID-19. For example, Atyeo *et al* suggest that strong early Ab responses to the N antigen are associated with high disease severity (21) and Ng *et al* suggest that cross-reactivity to the S2 domain of the spike antigen of common coronaviruses might offer some degree of protection from SARS-COV-2 infection (22). Multiplexing the N and spike antigens may also be useful for distinguishing natural infection from (spike-based) vaccination responses, as it would be expected that only infection would lead to the presence of antibodies reactive with the N antigen. However, caution should be used in solely relying upon the differential presence of anti-N and anti-spike Abs. Aside from individual variation, timing of collection may be important as some studies have shown that the duration of anti-N Abs is shorter than anti-spike Abs (23). In these cases, spike reactivity without N antigen reactivity might mistakenly be reported as a response to vaccination with a spike-based vaccine.

The MIA is readily adaptable to measuring responses of different Ig isotypes, different specimen matrices, and high throughput analyses. For example, our laboratory has used the MIA to detect IgM antibodies reactive with the SARS-CoV-2 N, RBD, and full-length spike in both newborn serum and adult CSF to suggest in situ infections ((24) and unpublished observations). However, isotype analyses may have their highest value in delineating the stage of infection. Although the full kinetics of antibody persistence, including isotypes, has yet to be determined, it might be expected that the presence of IgM would be indicative of a more recent infection, whereas IgG alone might indicate either a remote infection or a vaccine response. In terms of our own analyses of antibody isotypes at early points after the onset of infection, we noted the early onset of robust IgG production relative to a more canonical antibody induction pattern. The quick production of IgG has been observed by others (25-27). The two individuals shown in Figure 6 showed a rapid rise in IgG reactive with both the N and RBD antigens, relative to IgM and IgA. Both of those people were hospitalized and on ventilators at the time of blood collection. Our results are consistent with measurements on the same two individuals as reported by Yang *et al*, where IgM and IgG, also reactive with the N and RBD antigens, were assessed using a different assay system (Pylon COVID-19 IgM and IgG cyclic fluorescence assay) (27). That study examined a larger group and found that for most individuals, IgG was detected coincidentally or prior to IgM detection and that there was no significant difference associated with disease severity. This pattern, as illustrated by Figure 6, resembles a canonical secondary antibody response. As opposed to B cells, SARS-CoV-2 cross-reactive memory T cells, originating from prior infection with seasonal coronaviruses, have been identified (28). It is tempting to speculate that these cells help SARS-CoV-2-specific B cells and drive immediate antibody class switching. However, it is also possible that switch and heightened production of IgG is due to the general extensive inflammation caused by virus infection. It is also possible that the IgG antibodies produced are of higher affinity than the IgM and thus, may be more easily detectable in immunoassays.

Our purpose in developing the NYS COVID MIA was to engage in clinical testing for COVID Abs. To that end, we have tested both clinical specimens for assessment of patient responses to SARS-COV-2 and we have also tested more than 27,000 sera of potential donors of convalescent plasma. The assay has been further modified to detect Abs in dried blood spots and used in high throughput immunosurveillance studies to monitor SARS-CoV-2 exposure in New York State (29). The MIA continues to evolve through evaluation and possible addition of spike antigens from variants of concern. Further, for the convalescent plasma screening effort, we currently are aligning the MIA Index values with virus neutralization values to better identify “high titer” neutralizing plasma. Finally, our future efforts will include validation of the MIA using standardized sera, including the WHO International Standard (https://www.nibsc.org/documents/ifu/20-136.pdf), so that results can be harmonized and reported as quantitative Binding Antibody Units.

## 4. Materials and Methods

### 4.1. COVID-19 Serum Samples

Studies were performed on sera from deidentified specimens submitted to the New York State Department of Health in response to a broad request for assay development and validations. The request was initially made prior to and just following the first reported COVID-19 case in New York (March 1, 2020). The following institutions submitted serum specimens after RT-PCR confirmation of SARS-CoV-2 infection: Northwell Hospital Systems, University of Rochester Medical Center, Columbia University Medical Center, New York Presbyterian/Weill Cornell Medical Center, New York Presbyterian Hospital (NYPH)/Columbia University (CUMC) and Weill Cornell Medical Center (WCMC), and Mount Sinai Health System. These submitted specimens were “early acute” (collected 1-25 days post onset of symptoms). Some acute specimens (collected 1-25 days post onset of symptoms) were from COVID-19 hospitalized individuals at NYPH/WCMC. Additional specimens, obtained from the New York Blood Center, were sera obtained from healthy, convalescent patients who had recovered from COVID-19 >14 days prior to serum collection. These specimens were from individuals volunteering to be tested as potential donors of plasma for passive Ab transfer. All specimens were stored at 4°C until testing was completed (<1 week) and transferred to −80°C for long-term storage. Aliquots were made to minimize freeze-thaw. All testing and archiving of human specimens were done under a declared New York State Public Health Emergency and also approved by NYSDOH Institutional Review Board (IRB 20-021). Guinea pig antiserum reactive with SARS-CoV-1 was purchased from BEI Resources (Manassas, VA).

### 4.2 Non-COVID-19 Serum Samples

Specificity and control serum panels were deidentified specimens consisting of 1) an equal representation from presumed healthy individuals and received prior to 2019 that were obtained from (downstate New York) the New York Blood Center or (upstate New York) the American Red Cross; 2) positive identification for known pathogens or autoantigens and obtained through purchase, donation, or submission for clinical testing; and, 3) acute phase (non-COVID-19) respiratory infection obtained from NYPH/WCMC. For the last, where indicated, the agent of infection was identified using molecular testing.

### 4.3 Reagents

For MIAs, wash buffer and phosphate buffered saline pH 7.4, 0.05% sodium azide (PBS-TN) were purchased from Sigma (Sigma Aldrich, St. Louis, MO). Chemicals, 1-ethyl-3-(3-dimethylaminopropyl) carbodiimide hydrochloride (EDC) and N-hydroxysulfosuccinimide (sulfo-NHS), were supplied by Pierce Chemicals (Pierce, Rockford, IL). Microspheres, calibration microspheres, and sheath fluid were obtained from Luminex Corporation (Luminex Corp., Austin, TX). R-phycoerythrin (PE)-conjugated goat anti-human Ig, goat anti-human IgG, anti-human IgM, anti-human IgA, mouse anti-human IgG1, mouse anti-human IgG2, mouse anti-human IgG3, and mouse anti-human IgG4 were purchased (Southern Biotech). Recombinant SARS-CoV-2 nucleocapsid and spike S2 domain were purchased (Native Antigen, Oxfordshire, UK). Recombinant nucleocapsid and spike antigens from the common human coronaviruses (OC43, 229E, HKU-1, NL-63) were purchased (Native Antigen, Oxfordshire, UK). Recombinant SARS-CoV-2 RBD and spike S1 domain were provided by MassBiologics (Boston, MA), and produced as described below. Recombinant SARS-1 nucleocapsid was produced at the Wadsworth Center as described below. For some experiments, SARS CoV-2 RBD was also a kind gift from Dr. Florian Krammer, (Mount Sinai Health System). SARS-CoV-1 spike proteins were purchased from BEI Resources (Manassas, VA).

### 4.4 Recombinant Antigens

The amino-acid sequence of the SARS-CoV-2 S glycoprotein sequence (GeneBank: MN908947) were used to design a codon-optimized version for mammalian cell expression. The synthetic gene encoding the Receptor Binding Domain (RBD) a.a. 319-541 and S1 subunit a.a. 1-604 of the S glycoproteins were cloned into pcDNA 3.1 Myc/His in-frame with c-Myc and 6-histidine epitope tags that enabled detection and purification. The cloned genes were sequenced to confirm that no errors had accumulated during the cloning process. All constructs were transfected into Expi293 cells using ExpiFectamine 293 Transfection Kit (Thermo Fisher), and recombinant proteins were purified by immobilized metal chelate affinity chromatography using nickel-nitrilotriacetic acid (Ni-NTA) agarose beads. Proteins were eluted from the columns using 250 mmol/L imidazole and then dialyzed into phosphate-buffered saline (PBS), pH 7.2 and checked for size and purity by sodium dodecyl sulfate polyacrylamide gel electrophoresis (SDS-PAGE). For production of the recombinant SARS-CoV-1 Nucleocapsid protein, a cDNA copy of the N gene of SARS-CoV was produced by reverse transcription-PCR of RNA purified from Vero E6 cells that had been infected with SARS-CoV (strain Urbani; GenBank accession number AY278741). The N gene cDNA was ligated into the bacterial expression vector pET-28a(+) (Novagen (Merck-Millipore) Burlington,MA) for expression of carboxy-terminal His6-tagged full-length N protein. Following induction in RosettaTM BL21(DE3)pLysS E. coli cells (Novagen (Merck-Millipore) Burlington,MA), his-tagged N protein was purified by metal chelation chromatography on a Ni-NTA column (Novagen (Merck-Millipore) Burlington,MA). The purified N protein migrated in SDS-PAGE in accord with its predicted molecular mass of 47.1 kDa and was estimated to be greater than 98% pure.

### 4.5. Microsphere Immunoassay (MIA)

Specimens were assessed for the presence of reactive antibodies using an MIA modified from a previously described procedure (30). Briefly, recombinant antigens were covalently linked to the surface of fluorescent microspheres (Luminex Corporation, Austin, TX). Serum samples (25 μL at 1:100 dilution) and antigen-conjugated microspheres (25 μL at 5×10^4^ microspheres/mL) were mixed and incubated 30 minutes at 37°C before washing and further incubation with phycoerythrin (PE)-conjugated secondary antibody. The PE-conjugated antibodies were chosen to specifically recognize, as indicated, total antibodies (pan-Ig), or, individually IgM, IgA, IgG, IgG1, IgG2, IgG3, IgG4. After washing and final resuspension in buffer, the samples were analyzed on a FlexMap 3D analyzer using xPONENT software, version 4.3 (Luminex Corporation, Austin, TX).

### 4.6 High-throughput Instrument Detection of Antibodies to SARS-CoV-2

Specimens were assessed for the presence of reactive antibodies using: 1) the Ortho-Clinical Diagnostics (Rochester, NY) Vitros anti-SARS-CoV-2 IgG chemiluminescent immunoassay; 2) the Abbott Laboratories (Abbott Park, IL) SARS-CoV-2 IgG chemiluminescent microparticle immunoassay); 3) the bioMerieux (Durham, North Carolina) VIDAS SARS-COV-2 IgG immunoassay. The Abbott Laboratories SARS-CoV-2 IgG assay is a qualitative chemiluminescence test that we performed on the Abbott Architect i1000SR automated immunoassay analyzer. The target antigen is the SARS-CoV-2 N protein and only IgG antibodies were measured. Testing was performed in accordance with the manufacturer’s instructions using undiluted serum. The FDA EUA listed performance characteristics are: Sensitivity, 100% (95.8%-100%); Specificity, 99.6% (99.0% −99.9%). The Ortho-Clinical Diagnostics anti-SARS-CoV-2 IgG assay is a qualitative chemiluminescence test that we performed on the Ortho Vitros 5600 automated immunoassay analyzer. The target antigen is the S1 subunit of the SARS-CoV-2 spike protein and only IgG antibodies were measured. Testing was performed in accordance with the manufacturer’s instructions using undiluted serum. The FDA EUA listed performance characteristics are: Sensitivity, 90.0% (76.9% - 96.0%); Specificity, 100% (99.1% - 100%). The bioMerieux VIDAS SARS-COV-2 IgG immunoassay is a qualitative enzyme-linked fluorescence assay that we performed on the Vidas-3 automated analyzer. The target antigen is the RBD of the SARS-CoV-2 spike protein and only IgG antibodies were measured. Testing was performed in accordance with the manufacturer’s instructions using undiluted serum. The FDA EUA listed performance characteristics are: Sensitivity, 100% (88.3 - 100%); Specificity, 99.9% (99.4% - 100%).

### 4.7 Reporting of Statistical Methods

All graphs and statistical analyses to determine Spearman’s rank correlation coefficients were done using Prism 9.0 (Graphpad, San Diego, CA). Spearman’s correlations were calculated using all complete pairs of variables in the dataset.

## Data Availability

All data generated or analysed during this study are included in this submitted article (and its supplementary information files)

## 6. Acknowledgments

The authors would like to acknowledge and thank the following Wadsworth Center laboratory members for their expert technical assistance: June Chan, Margaux Hales, Colleen Walsh, Geraldine Quinones, Brian Grimwood. We thank Dr. Lili Kuo her advice and insights on the expression of viral proteins. We especially thank Drs. Susan Wong and Paul Masters for their helpful advice and their critical reading of this manuscript and Dr. Steven Spitalnik for support and advice. We would like to acknowledge the Wadsworth Center core facilities that contributed to this work: WC Tissue Culture and Media Core and the WC Sequencing Core.

## Footnotes

### ^1^Abbreviations

COVID-19: coronavirus disease 2019
SARS-CoV-2: Severe acute respiratory syndrome coronavirus 2
HuCOV: human endemic (seasonal) coronaviruses
Ab: antibody
DIL: Diagnostic Immunology Laboratory at the Wadsworth Center
N: nucleocapsid protein
RBD: receptor-binding domain
MFI: median fluorescent intensity
FDA: Food and Drug Administration
EUA: Emergency Use Authorization
MIA: microsphere immunoassay

