## Supplementary Information for "COVID-19 Serology in New York State Using a Multiplex Microsphere Immunoassay"

**Supplementary Figure Legends**

Supplementary Figure 1

**Antibodies to common huCoV's in healthy donor sera.** Nucleocapsid (*A*) and Spike antigens (*B*) from common human coronaviruses (OC43, HKU1, 229E, NL63) or SARS-CoV-2 were conjugated to microspheres and used in the MIA to determine binding with sera from pre-COVID-19 healthy blood donors. Shown is the median fluorescence intensity of total bound antibodies detected with PE-anti-human Ig, along with the 3 SD and 6 SD cutoff values determined for SARS-CoV-2 (used in the clinical NYS CoV MIA).

Supplementary Figure 2

**Relative binding to SARS spike antigens in individual COVID-19 sera.** The full spike antigens from SARS-CoV-1 (*circles*) or SARS-CoV-2 (*squares*), and the SARS CoV-2 spike components: RBD (*upward triangles*), S1 (*downward triangles*), and S2 (*diamonds*), were conjugated to microspheres and used in the MIA to determine binding of total antibodies in sera from COVID-19 convalescent individuals. Results are expressed as index values relative to cutoff MFI's plus 3 SD, determined for each individual antigen. Shown are results from 8 individual patients (part of the specimens used to determine values in Figure 4B).

Supplementary Figure 3

**Production of different antibody classes reactive with SARS-CoV-2 target antigens**. Serum specimens from individuals with SARS-CoV-2 infections, confirmed using RT-PCR, and reported symptom onset dates, were assessed for total antibody (Ig), IgM, IgG, and IgA antibodies to different SARS-CoV-2 target antigens (Nucleocapsid, full spike, and separate spike components (RBD, S1 subunit, S2 subunit). Shown are individual index values for each class to the individual target antigens.

Supplementary Figure 4

**Production of different IgG subclasses reactive with SARS-CoV-2 target antigens in the MIA**. Serum specimens from individuals with SARS-CoV-2 infections, confirmed using RT-PCR, and reported symptom onset dates were assessed for IgG1, IgG2, IgG3, and IgG4 antibodies to SARS-CoV-2 nucleocapsid, spike antigen and spike components: RBD, S1, and S2 target antigens. Shown are relative amounts of antibodies based upon Index values along with the 3 SD (Indeterminate threshold) and 6 SD cutoff (Positive threshold) values.

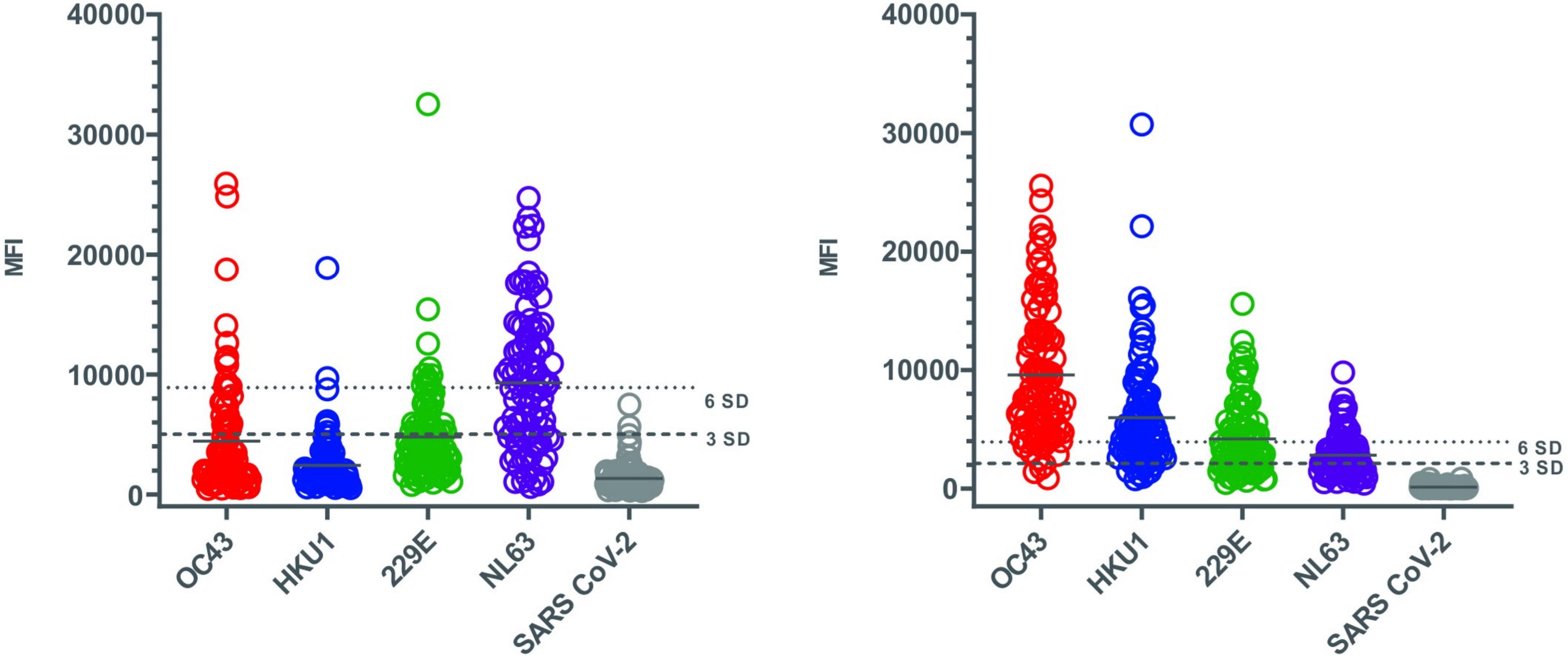

**Supplementary Figure 1**

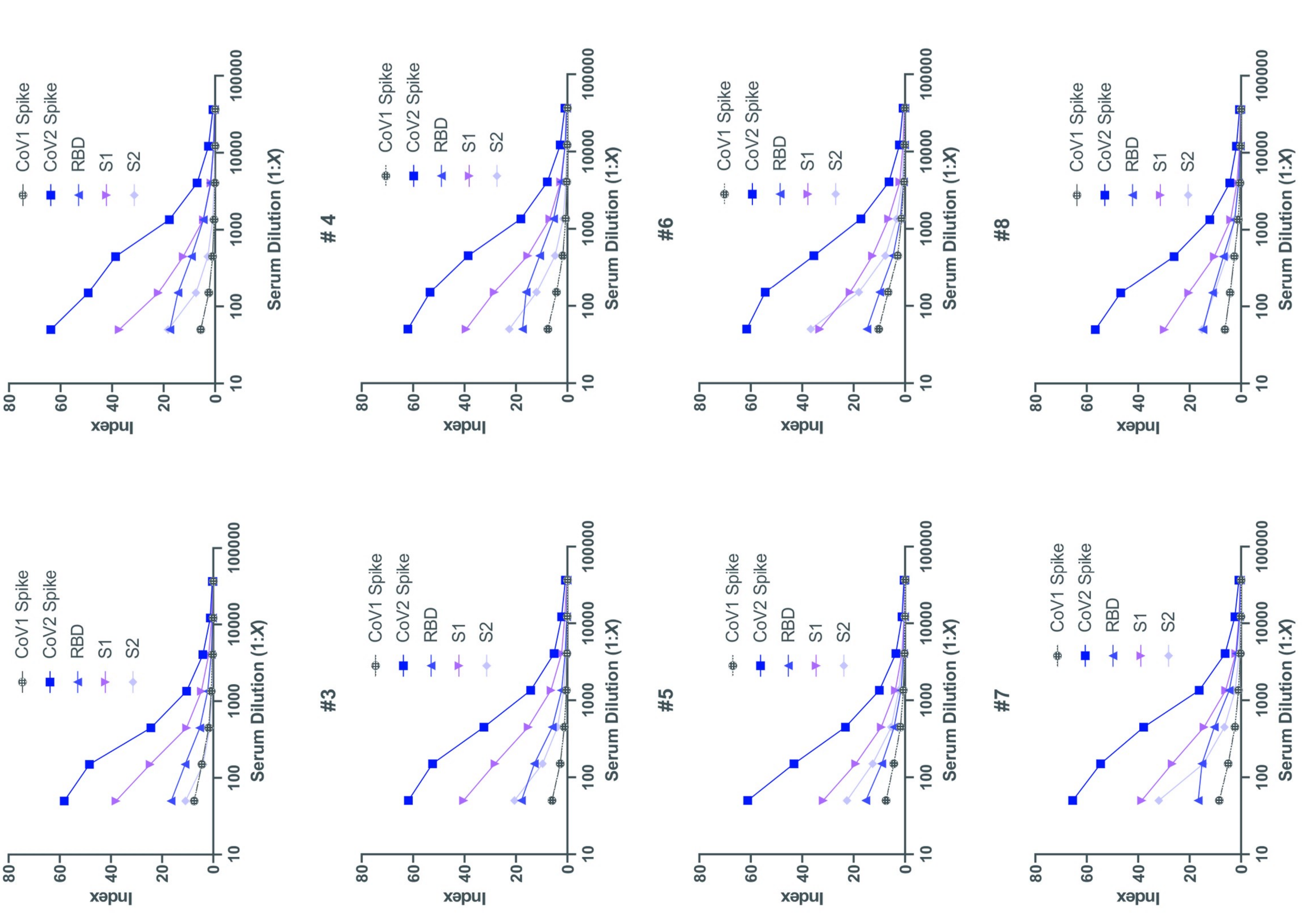

**Supplementary Figure 2**

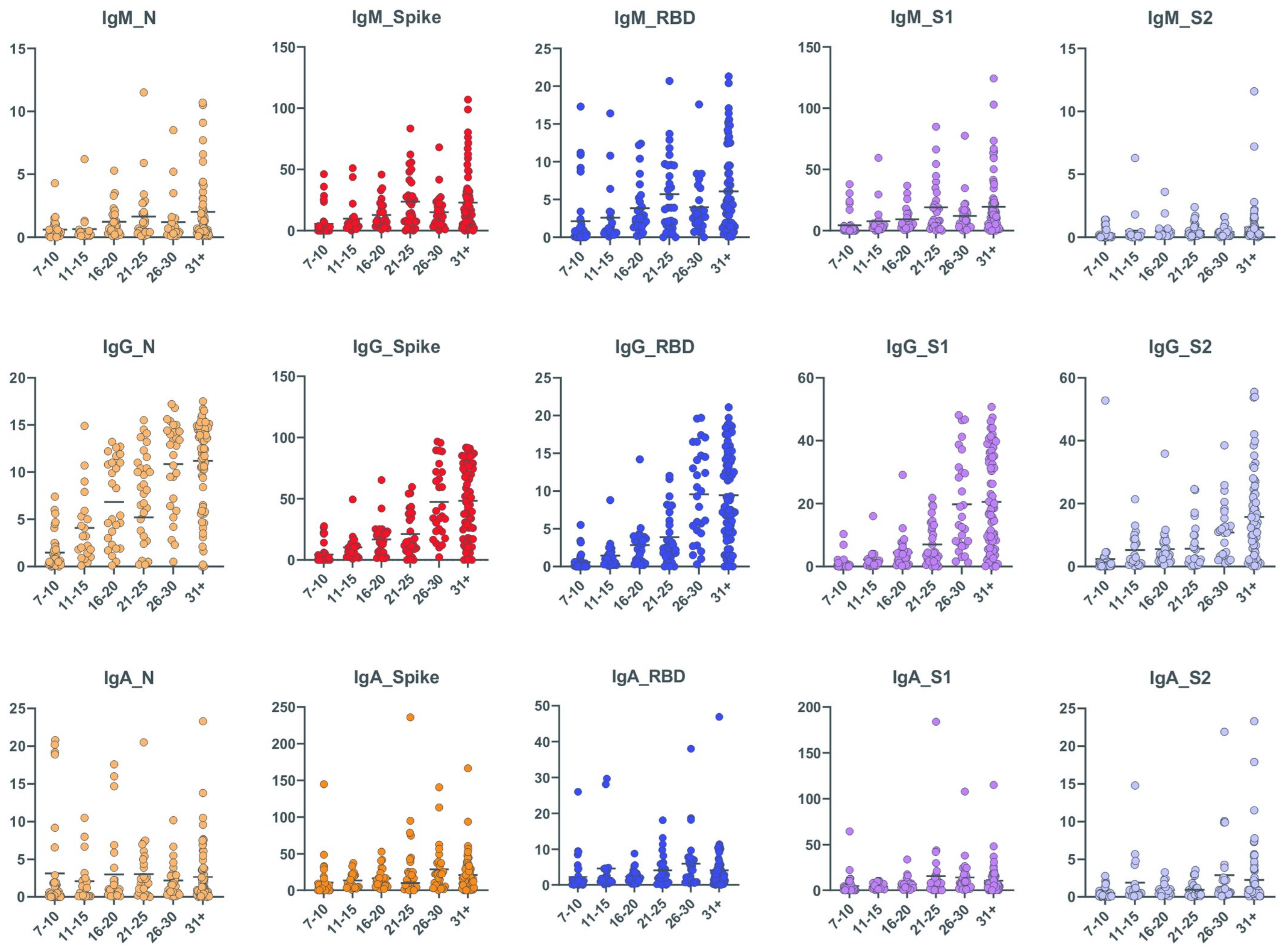

**Supplementary Figure 3**

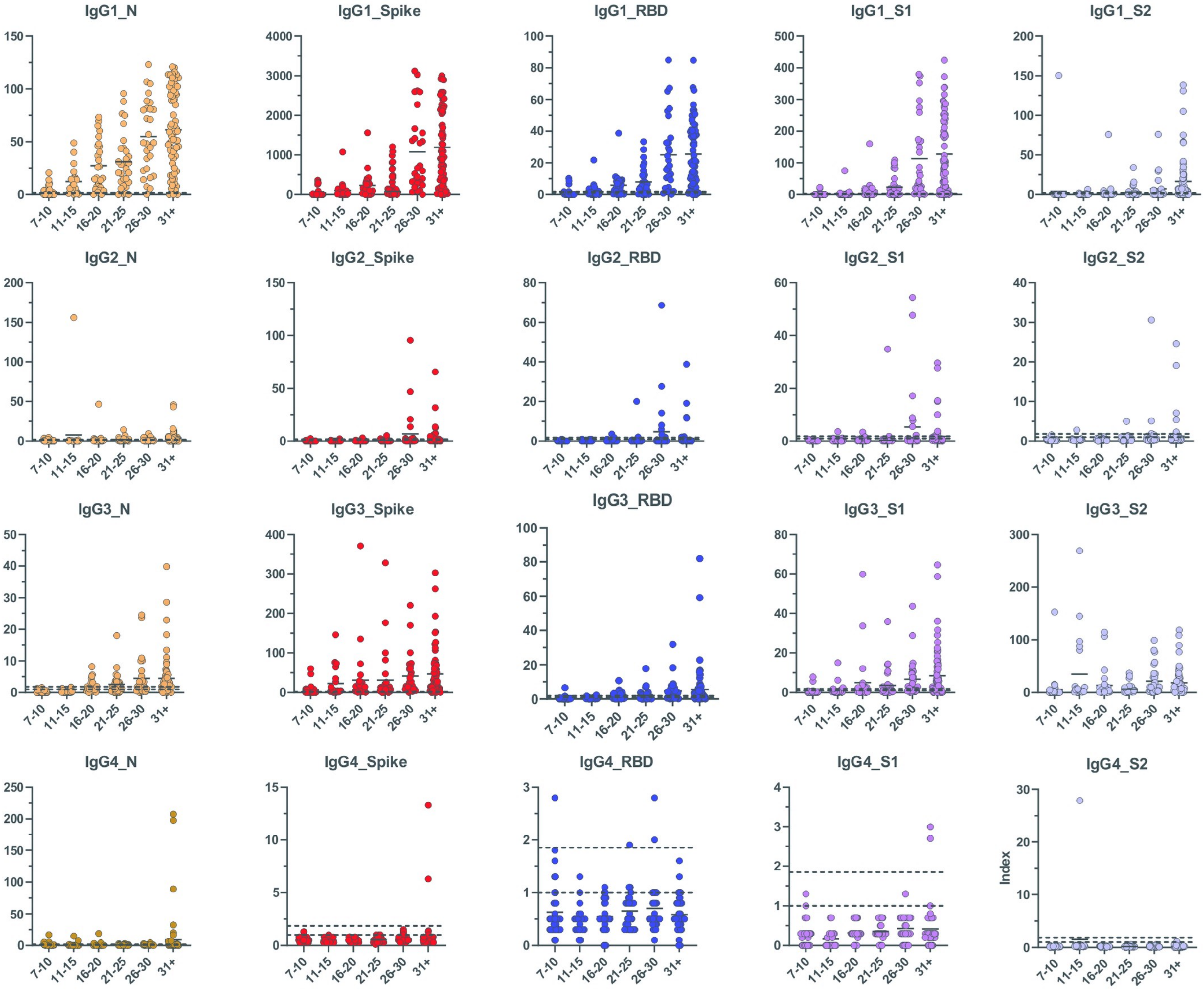

**Supplementary Figure 4**

| | Assay | MIA | MIA | MIA | MIA | MIA | VIDAS | Ortho | Abbott |
|---|---|---|---|---|---|---|---|---|---|
| | **Antigens** | **N/RBD** | **N/RBD/Spike** | **N** | **RBD** | **Spike** | **RBD** | **S1** | **N** |
| **Pre-COVID** | Positive | 1 | 2 | 0 | 1 | 2 | 1 | 0 | 0 |
| | Indeterminate | 5 | 6 | 2 | 3 | 2 | 0 | 0 | 0 |
| | Negative | 94 | 92 | 98 | 96 | 96 | 99 | 100 | 100 |
| | *Reactivity (%)* | *1* | *2* | *0* | *1* | *2* | *1* | *0* | *0* |
| | *Specificity (6SD*)(%)* | *99* | *98* | *100* | *99* | *98* | *99* | *100* | *100* |
| | *Specificity (3SD*)(%)* | *94* | *92* | *98* | *96* | *96* | | | |
| **Convalescent (excluding MIA Indeterminates and Negatives)** | Positive | 300 | 300 | 300 | 288 | 298 | 294 | 296 | 291 |
| | Indeterminate | 0 | 0 | 0 | 9 | 0 | 0 | 0 | 0 |
| | Negative | 0 | 0 | 0 | 3 | 2 | 6 | 4 | 9 |
| | *Sensitivity (6SD*) (%)* | *100* | *100* | *100* | *96* | *99* | *74* | *98.7* | *97* |
| | *Sensitivity (3SD*) (%)* | *100* | *100* | *100* | *99* | *99* | | | |
| **Convalescent that includes MIA Indeterminates and Negatives** | Positive | 352 | 390 | 344 | 319 | 388 | 328 | 361 | 295 |
| | Indeterminate | 32 | 0 | 28 | 42 | 0 | 0 | 0 | 0 |
| | Negative | 16 | 10 | 28 | 39 | 12 | 72 | 39 | 105 |
| | *Sensitivity (6SD*) (%)* | *88* | *97.5* | *86* | *79.8* | *97* | *82* | *90.2* | *73.8* |
| | *Sensitivity (3SD*) (%)* | *96* | *97.5* | *93* | *90.2* | *97* | | | |

**Supplementary Table 1. Comparison of the NYS MIA with three commercial tests.** 100 Pre-COVID serum specimens (*top panel*) and 400 serum specimens, collected from COVID-19 convalescent individuals 4-8 weeks post symptom onset, were screened using the NYS MIA (N plus RBD) and then re-tested using the NYS MIA (N plus RBD plus Spike) and three commercial tests on instrument platforms ((Biomerieux VIDAS, Ortho Vitros, Abbott Architect). One hundred specimens that screened as indeterminate or negative were excluded from the initial analysis (*middle panel*) and then included (*bottom panel*) to show test performance at the lower range of antibody concentration. All assays, including the MIA, specifically measured IgG antibodies. *Specificity and Sensitivity at different SD thresholds are for NYS MIA only.

| Days from onset | Samples tested (N) | RT-PCR result | NY SARS-CoV MIA Result as Compared to PCR | | | Sensitivity (95%CI) |
|---|---|---|---|---|---|---|
| | | | Positive | Ind. | Negative | |
| **IgM** | | | | | | |
| **7 – 15** | 48 | Pos | 10 | 8 | 30 | **10/48**<br>**20.8% (11.73% - 34.26%)** |
| **16 – 25** | 17 | Pos | 11 | 1 | 5 | **11/17**<br>**64.7% (41.30% - 82.69%)** |
| **26 – 30** | 28 | Pos | 21 | 2 | 5 | **21/28**<br>**75% (56.64% - 87.32%)** |
| *Total* | *93* | | | | | |
| **IgG** | | | | | | |
| **7 – 15** | 48 | Pos | 16 | 5 | 27 | **16/48**<br>**33.3% (21.68% - 47.46%)** |
| **16 – 25** | 17 | Pos | 12 | 1 | 4 | **12/17**<br>**70.6% (46.87% - 86.72)** |
| **26 – 30** | 28 | Pos | 28 | 0 | 0 | **28/28**<br>**100% (87.94% - 100%)** |
| *Total* | *93* | | | | | |
| **IgA** | | | | | | |
| **7 – 15** | 48 | Pos | 28 | 5 | 15 | **28/48**<br>**58.3% (44.28% - 71.15%)** |
| **16 – 25** | 17 | Pos | 12 | 0 | 5 | **12/17**<br>**70.6% (46.87% - 86.72%)** |
| **26 – 30** | 28 | Pos | 21 | 2 | 5 | **21/28**<br>**75% (56.64% - 87.32%)** |
| *Total* | *93* | | | | | |

**Supplementary Table 2. Determination of MIA Sensitivity.** A total of 93 serum specimens from individuals with SARS-CoV-2 infections, confirmed using RT-PCR, and reported symptom onset dates were assessed for antibodies of the indicated isotypes to the SARS-CoV-2 Nucleocapsid antigen and the RBD. A positive specimen is ≥6 SD above the cutoff for either one of the target antigens; an indeterminate is ≥3 SD above the cutoff for either or both of the target antigens; a negative is a sample with no reactivity to either target antigen. Sensitivities at different periods post symptom onset are shown on the right as the number of MIA positive specimens/total specimens tested.
